# The Target ALS Global Natural History Study: Cross-platform proteomics to accelerate biofluid biomarker and drug target discovery in amyotrophic lateral sclerosis

**DOI:** 10.64898/2026.06.13.26355379

**Authors:** Daisuke Yasui, Daniel Weatherill, Laura Dugom, Sophia Weiner, Lathika Gopalakrishnan, Huy Tran, Björn Oskarsson, Kyra Nagle, Timothy Miller, Gilbert Gutierrez, John Ravits, Benjamin Hoover, Matthew Harms, Neil Shneider, Lizzi Neylon, Whitney Dailey, Shafeeq Ladha, Cassandra Holmes, Joseph Lee, Nicholas Streicher, Shakti Nayar, Brent T. Harris, Manish Raisinghani, Henrik Zetterberg, Johan Gobom, Amy Easton, Robert Bowser, Cindy V. Ly

## Abstract

Amyotrophic lateral sclerosis (ALS) is a fatal, rapidly progressive neurodegenerative disease of motor neurons for which therapeutics are limited. Improved biomarkers are imperative to improve patient care and therapeutic development. Here, we employed 35-plex isobaric tandem mass tag labeling based on isobutyl-proline reporter group (TMTpro) to perform unbiased proteomic analysis of cerebrospinal fluid (CSF) and plasma from control (n= 28, n= 31) and sporadic ALS (sALS) (n= 39, n= 41), from the Target ALS Global Natural History Study (TALS GNHS). We identified 2,875 proteins in CSF and 1,118 proteins in plasma and identified known and novel differentially expressed proteins (DEPs) between controls and sALS, some of which were orthogonally validated using immunoassay. Comparison of TMTpro-MS and Olink proximity extension assay proteomics revealed common and non-overlapping differentially expressed proteins illustrating strengths unique to each platform. This initial cross-sectional proteomic study of biofluids from the TALS GNHS, with unrestricted availability of study results to the research community, highlights the potential of this resource as a potent platform for ALS biomarker discovery.

## Introduction

Amyotrophic lateral sclerosis (ALS) is a devastating neurodegenerative disorder characterized by motor neuron loss that leads to weakness, muscle atrophy, respiratory failure, and death within 2-5 years of symptom onset. Only about 10% of ALS cases are due to genetic etiology, with the remaining 90% of cases considered sporadic or singleton ALS (sALS) without a known cause (Feldman, 2022 ^1^). ALS is a clinically and genetically heterogeneous disorder that affects patients globally. Advancing effective therapeutics for this condition relies on discovery of novel biomarkers to improve early disease detection, monitor disease progression, and determine pharmacodynamic response in clinical trials. ALS is a mechanistically complex disorder so unique biomarkers representing various facets of disease pathogenesis are also desired as the future therapeutic pipeline encompasses a diversity of targets aimed at multiple facets of disease. Tofersen (i.e. Qalsody); the recently approved treatment for disease caused by mutations in superoxide dismutase 1 (*SOD1*) familial ALS (fALS), by demonstrating lowered neurofilament levels months before demonstration of clinical stabilization and improved survival *SOD1* ALS constitutes ∼20% of fALS but only ∼2% of the ALS population^2^ (Miller 2025). The accelerated approval of tofersen based on the reduction of a biomarker of neurodegeneration has underscored the importance of having robust biofluid biomarkers in clinical trial design.

Although neurofilament light chain (NfL) and phosphorylated neurofilament heavy chain (pNfH) have emerged as informative proxies of neurodegeneration, it is clear that neurofilament proteins can only serve as biomarkers in specific contexts of use but not all. Neurofilament proteins are non-specific biomarkers that are elevated in numerous neurological conditions^3^. Neurofilament release reflects a downstream stage of neurodegeneration, axonal and neuronal cell death, and signals damage once it has already occurred. Biomarkers that capture disease activity at earlier stages, prior to cell death would therefore be advantageous, particularly for early intervention. Further, neurofilament proteins have a long half-life and therefore may not be ideal for early detection of drug treatment effects. Protein biomarkers that may be used separately or in conjunction with neurofilaments have been identified. For example, inflammatory markers have been shown to be differentially expressed in ALS. Chitinases are elevated in CSF from ALS patients compared to control^4,5,6^, T-regulatory cell number is reduced and correlates with ALS disease progression^7^, and p75 is increased in urine from ALS and rises during disease progression^8^. Interestingly, tau proteins phosphorylated at amino acid 181 and/or 217 are both elevated in blood of patients with ALS and have been associated with muscle fiber atrophy^9,10^. Troponin T protein that is derived from muscle is increased in the blood of ALS patients and increases over time within patients^11^. In addition, a recent study reported at least thirty differentially expressed proteins linked to muscle function, nerves, and energy metabolism^12^. Notably, the majority of ALS cases (∼97%) are characterized by TAR DNA-binding protein 43 (TDP-43) pathology. TDP-43 is an RNA-binding protein that, in a disease context, mislocalizes from the nucleus to the cytoplasm and forms pathological aggregates^13^. The loss of nuclear function has been associated with altered mRNA splicing leading to aberrant translation of cryptic peptides that lead to protein loss-of-function (e.g., UNC13, STMN2, KCNQ2) or neoantigens (i.e. HDGFL2) some of which can be detected in CSF^14,15,16,17,18^. While these advances are promising, none of these peptides yet meet criteria for advancing into clinical grade biomarker assays. There remains a need both for biomarkers that reflect disease specificity and clinical heterogeneity.

The Target ALS Global Natural History Study (TALS GNHS) was launched in 2021 as an effort to collect longitudinal biofluids including cerebrospinal fluid (CSF) and blood paired with comprehensive clinical, genetic, and molecular data from a diverse geographic and ethnic representation of ALS participants to provide a global platform for biomarker discovery informed by considerations of regional disease heterogeneity. In addition, data collected is publicly available to accelerate these efforts.

Here we conducted unbiased proteomic analysis using state of the art 35-plex TMTpro-MS as well as antibody-based proteomics using Olink HT platform on CSF and plasma from controls and sALS, from the TALS GNHS biorepository in a cross-sectional study and performed orthogonal validation of selected top proteomic hits using immunoassays in a separate sample cohort.

## Materials and Methods

### The Target ALS Global Natural History Study (TALS GNHS) Cohort

The TALS GNHS includes participants with ALS, healthy controls, and asymptomatic mutation carriers. CSF and plasma were collected longitudinally from participants with sALS or fALS approximately every 4 months as well as healthy controls or asymptomatic carriers of mutations conferring ALS risk once a year. All biofluid samples were collected after obtaining patient informed consent and as part of approved institutional review board (IRB) protocols at individual academic sites involved in the TALS GNHS (NCT05137665). We examined CSF from healthy control (n= 28) and sALS (n= 39), as well as plasma from healthy control (n= 31) and sALS (n= 41) in a cross-sectional study focused on early visit 1 or visit 2 timepoints after study enrollment. Biosamples are linked to detailed clinical information including age, gender, site of onset, and ALS Functional Rating Scale-Revised (ALSFRS-R).

Data can be assessed through the Target ALS Data Engine (https://dataengine.targetals.org/collections).

### Validation Cohort

An additional biofluid cohort was provided by the Washington University (WashU) ALS biorepository that included CSF from healthy control (n= 10) and sALS (n= 15) as well as plasma from healthy control (n= 10) and sALS (n= 20). Characteristics of the WashU cohort are detailed in Table 12 for CSF and Table 13 for plasma. Biofluids were collected as part of ongoing research protocols approved by the Washington University institutional review board (IRB) with appropriate informed consent.

### Protein immunodepletion, reduction, alkylation, digestion and Tandem Mass Tag (TMTpro) 35-plex labelling of biofluids

Cerebrospinal fluid (CSF) and plasma samples were processed in parallel using identical workflows as previously described^19^. Briefly, high-abundance proteins were depleted from each sample using the High-Select Top14 resin (Thermo Scientific) according to the manufacturer’s instructions. Following depletion, cysteine disulfide bonds were reduced and free thiols carbamidomethylated by incubation with iodoacetamide (10mM). Proteins were then digested in solution with a mixture of sequencing-grade Trypsin and Lys-C proteases (Promega). Resulting peptides were labeled with TMTpro 35-plex reagents (Thermo Scientific), combined into multiplexed sets, and desalted by solid-phase extraction (SPE) performed with reversed-phase C_18_ cartridges (Sep-Pak C18 light) using a vacuum manifold. Each TMT set was further fractionated by reversed-phase HPLC under high-pH conditions using an UltiMate 3000 LC System (Thermo Scientific) into 24 concatenated peptide fractions, to increase proteome coverage as previously described^20^.

### LC-MS/MS Acquisition

Peptide fractions were analyzed by LC-MS on a Vanquish Neo Nano-LC system coupled to a Tribrid Eclipse Orbitrap mass spectrometer, equipped with a Easy Spray ion source and a FAIMS Pro Duo interface (all from Thermo Scientific). The LC was operated in trap-and-elute mode, using a PepMap C18 trap column (100 Å, 300 μm x 5 mm) and an Easy Spray PepMap Neo C18 analytical column (75 μm x 500 mm). Peptides were separated using a linear gradient of Buffer A (0.1% formic acid) Buffer B (80% acetonitrile, 0.1% formic acid), increasing from 4% to 10% Buffer B over 1 min, followed by an increase from 10% to 40% Buffer B over 68 min. The mass spectrometer was operated in data-dependent acquisition (DDA) mode, acquiring full MS1 scans in the Orbitrap at a resolving power of 120,000 (m/z 200) followed by higher-energy collisional dissociation (HCD) MS2*MS2* scans at a resolving power of 75,000, with a 1.5-s cycle time. Two compensation voltages (CVs; -45V and -65V) were applied in alternating fashion to enhance gas-phase selectivity and suppress singly-charged non-peptide derived ions.

### Data Processing and Protein Identification

Raw data were processed using Proteome Discoverer 3.2 (Thermo Scientific). Peptide spectral matching was performed with the SequestHT search engine against the human subset of the UniProt/Swiss-Prot database. Search parameters permitted up to two missed tryptic cleavages and employed semi-specific cleavage specificity. Mass tolerances were set to ±5 ppm for precursor ions and 0.05 m/z for fragment ions (MS2). Fixed modifications included TMTpro labeling on peptide N-termini and lysine residues, as well as carbamidomethylation of cysteines. Peptide-level validation was carried out with Percolator, applying a false discovery rate (FDR) threshold of p < 0.01.

### Quantification and Normalization

TMT reporter-ion intensities were extracted in Proteome Discoverer and used for quantitative analysis. A pooled CSF/plasma reference (non-diagnosis specific) was included in each TMT set to facilitate cross-run normalization. For each sample channel, reporter ratios were calculated relative to the global pool and normalized to the median intensity of all quantified peptides/proteins within that channel. This workflow ensured consistent, high-precision relative quantification across all CSF and plasma samples.

### Antibody-based Proteomic Profiling

CSF and plasma samples were analyzed using Olink Explore HT (ThermoFisher). This platform uses affinity-based proximity extension assay technology (PEA) that employ pairs of antibodies that are each conjugated to a single-stranded DNA reporter probe. Upon antibody binding to target proteins, DNA probes hybridize to form a protein-specific barcode that is amplified and quantified on a NovaSeq 6000 (Illumina)^21^. Raw BCL files were converted into normalized protein expression (NPX) values presented on a log2 scale using a manufacturer-specified quality control and normalization process. As a result, out of the 5,416 proteins detected, 5,401 proteins were included in the analysis for CSF while all 5,416 detected proteins were included in the analysis for plasma.

### Validation of biomarker candidates via ELISA

Commercially available enzyme-linked immunosorbent assays (ELISA) were obtained for targets identified by 35plex TMTpro-MS to be differentially expressed in ALS CSF or plasma. For this purpose, we acquired ELISAs for SERPINA3 (ThermoFisher, H411RB), CKM (Abcam, ab264617), CHAD (LS Bio, LS-F11130). Assays were performed as per manufacturer’s protocols.

### Statistical Analysis

Cross-sectional analyses were performed on proteomics data derived using biofluids from the earliest single time point per individual constituting visit 1 or visit 2. Following normalization and quality control of the samples, raw abundance values were log_2_-transformed. Proteins that were not detectable in at least 50% of samples within any comparison group were excluded from analysis. Remaining missing values were retained as missing and were not imputed prior to statistical analysis. Pairwise comparisons between disease and control groups were conducted using Welch’s t-test to account for unequal variances. Multiple testing correction was applied using the Benjamini–Hochberg procedure (false discovery rate, FDR; α = 0.05). Statistical significance was defined as an adjusted p-value < 0.05. GraphPad Prism (Version 10.0.2) was used to generate volcano plots, box plots, and conduct statistical analysis. Spearman correlation analyses were carried out to determine relationships between Simoa NfL and relative protein abundances with clinical measures (p values < 0.05 were considered significant).

### Pathway Enrichment Analysis

Proteins meeting the significance threshold (adjusted p-value < 0.05) were stratified as increased or decreased relative to the comparison group. Each protein set was subjected to gene ontology (GO) enrichment analysis using Enrichr^22,23,24^. The input target lists were queried against the GO Biological Process 2025, GO Cellular Component 2025, and GO Molecular Function 2025 databases.

### Tissue-based classification of CSF and plasma DEPs

Differentially expressed proteins (DEPs) in sporadic ALS (sALS) versus healthy controls (HC) were classified by their annotated biological processes, grouped according to associated tissue and organ systems. The pipeline was applied separately to each combination of biofluid (CSF, plasma), regulation direction (up/down), and platform (35-plex TMTpro-MS, Olink HT). Gene symbols were queried against the MyGene.info API^25–27^ (v3, https://mygene.info/) to retrieve human Gene Ontology Biological Process (GO BP) annotations.

Each protein was assigned to a single category by keyword matching against its GO BP term descriptions, evaluating three neuromuscular categories in priority order: muscle/atrophy ("muscle", "myofibril", "sarcomere", "myosin", "actin", "motor neuron"), peripheral nerve ("peripheral", "Schwann", "myelin", "node of Ranvier", "neuromuscular"), and CNS ("brain", "spinal", "neuron", "axon", "synapse", "astrocyte", "microglia", "glial", and related terms); proteins matching more than one were assigned to the highest-priority category. Remaining proteins were labelled "Other" and screened against a secondary organ panel (heart/cardiovascular, liver, kidney, lung, bone/skeletal, immune/blood, skin/epithelial, digestive), taking the first match. For each bar chart, the three most frequent secondary organ categories were retained and the rest grouped as "Remaining Other." Per platform counts were normalized to 100% and shown as stacked bar charts annotated with total DEP counts. Analyses used Python (requests, pandas, matplotlib).

## Results

### Characteristics of participant cohorts

CSF from healthy control (n= 28) and sALS (n= 39) as well as plasma from the same and several additional participants (healthy control, n= 31; sALS, n= 41) were obtained from the TALS GNHS for cross-sectional proteomic analysis. Participants were well clinically characterized with regards to demographic and disease-specific clinical measures (Supp Tables 1-2).

### Accessing TALS GNHS Proteomic Data

Comprehensive de-identified proteomic datasets from the TALS GNHS Data Engine can be accessed without restriction by researchers worldwide by going to the Target ALS Data Engine (https://dataengine.targetals.org/collections) and navigating to the AGRI Global Natural History Study featured collection. Initial users will be prompted to apply for access to the data portal. Once access is approved, users will be able to explore and access proteomic and other multi-modal data (DNA Stack) and analyze the data within a secure, cloud-transparent framework (Verily Workbench). Detailed instructions for navigating the Target ALS Data Engine can found in a provided user manual (https://storage.googleapis.com/target-als-data-portal-public/user-manual/2024-03-28/TargetALS_DataPortal_UserManual.pdf).

### Proteomic analysis of differentially expressed proteins in sALS CSF

Unbiased quantitative proteomics of CSF was performed using 35-plex TMTpro-MS workflow (Figure 1 A). We identified a total of 2,875 proteins across 7 TMT sets (Supp. Figure 1 A, Supp. Table 5). After quality control to include CSF samples with at least 50% of proteins having quantification, 2,033-2,218 proteins among our ALS and healthy control cohorts were included for comparative analysis (Supp. Figure 1 B). We observed 73 proteins that were only detected in healthy controls and 30 in sALS that were excluded from analysis (Supp. Table 5).

**Figure 1.**
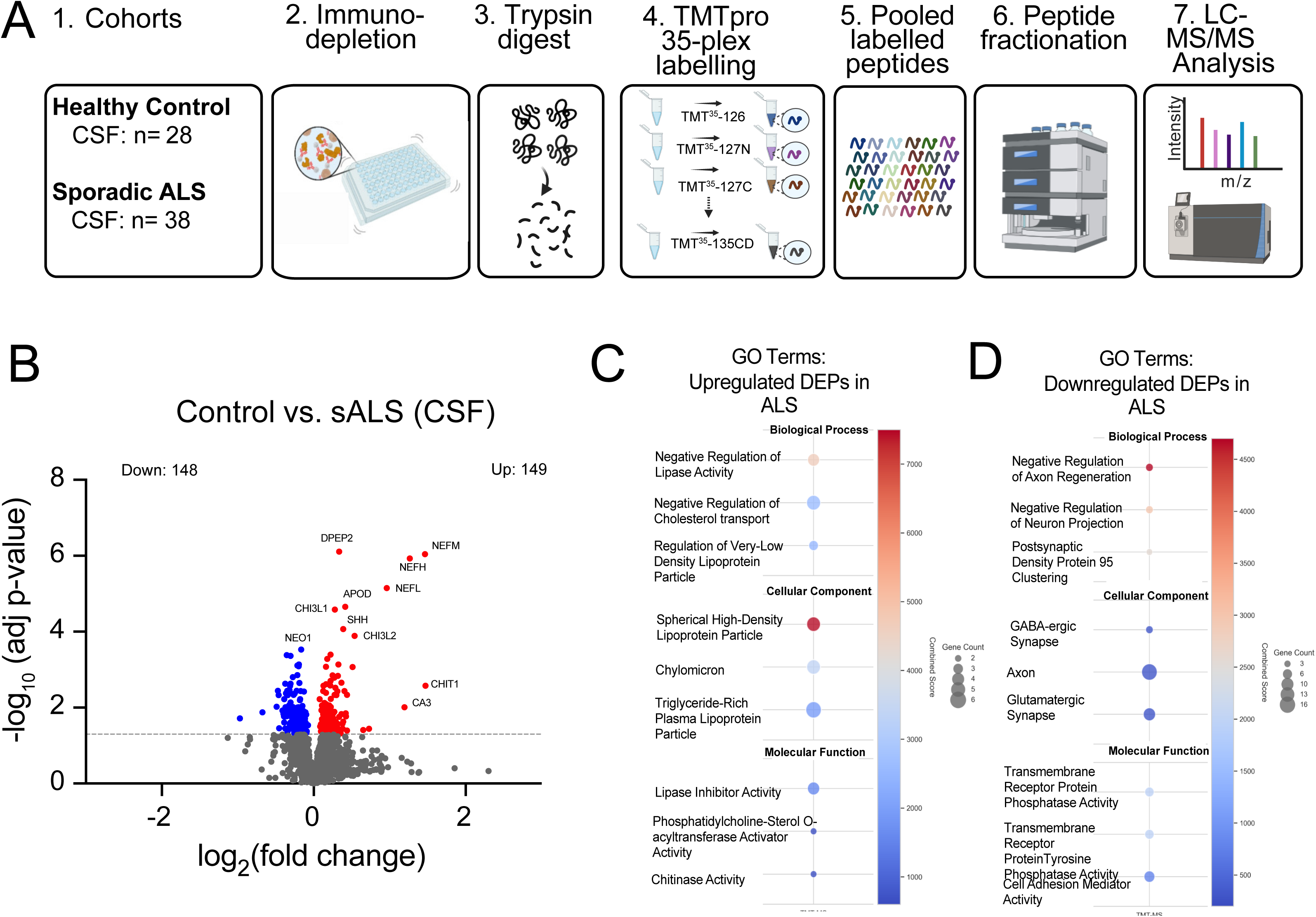
Overview of 35-plex TMTpro-MS methodology for CSF and differentially expressed proteins between control and ALS subtypes. Flow diagram of A) proteomic workflow used to determine proteomic differences in CSF from (1) control versus sporadic ALS (sALS). CSF is (2) immune-depleted to remove abundant proteins, (3) enzymatically digested, (4) undergoes TMT-labelling, and (4) samples are pooled prior to (5) analysis by liquid chromatography tandem mass spectrometry (LC-MS/MS). Volcano plot showing differentially expressed CSF proteins between B) controls (n= 28) versus sALS (n= 38). Adjusted p-values were derived from Benjamini-Hochberg correction for multiple comparisons. - log_10_(adj p-values) were plotted against log_2_(fold change). Significantly (adj p-value < 0.05) increased (red dot), decreased (blue dot), or not significantly changed proteins (gray dot) in ALS compared to control are indicated. Functional enrichment analysis of C) upregulated and D) downregulated genes in ALS compared to control using Enrichr analysis ^23^ shows associated gene ontology (GO) terms with combined score and number of linked proteins.

A total of 149 proteins were increased and 148 proteins decreased in sALS compared with healthy controls. Previously established ALS biomarkers neurofilament light (NEFL), neurofilament medium (NEFM), and neurofilament heavy (NEFH) chain as well as chitinase-3-like protein 1 (CHI3L1), chitinase-3-like protein 2 (CHI3L2), and chitinase-1 (CHIT1) were among the most significant DEPs indicative of neurodegeneration and inflammatory activation known to occur with disease ^3,4,5,6^ (Figure 1 B, Supp. Table 6). In addition, gene ontology suggests an enrichment in proteins involved in lipid regulation and cholesterol transport and downregulation of proteins involved in axonal regeneration and axon guidance and synaptic maintenance (Figure 1 C-D, Supp. Table 7). Novel DEPs included dipeptidase-2 (DPEP2), apolipoprotein-D (APOD), sonic hedgehog (SHH), and serpin family A member 3 (SERPINA3) which were increased as well as neogenin-1 (NEO1) which was decreased relative to healthy controls (Figure 2 A-J). APOD and SERPINA3 were previously shown to be at higher levels in the CSF of fast progressing ALS patients when compared to slow progressors^28^.

**Figure 2.**
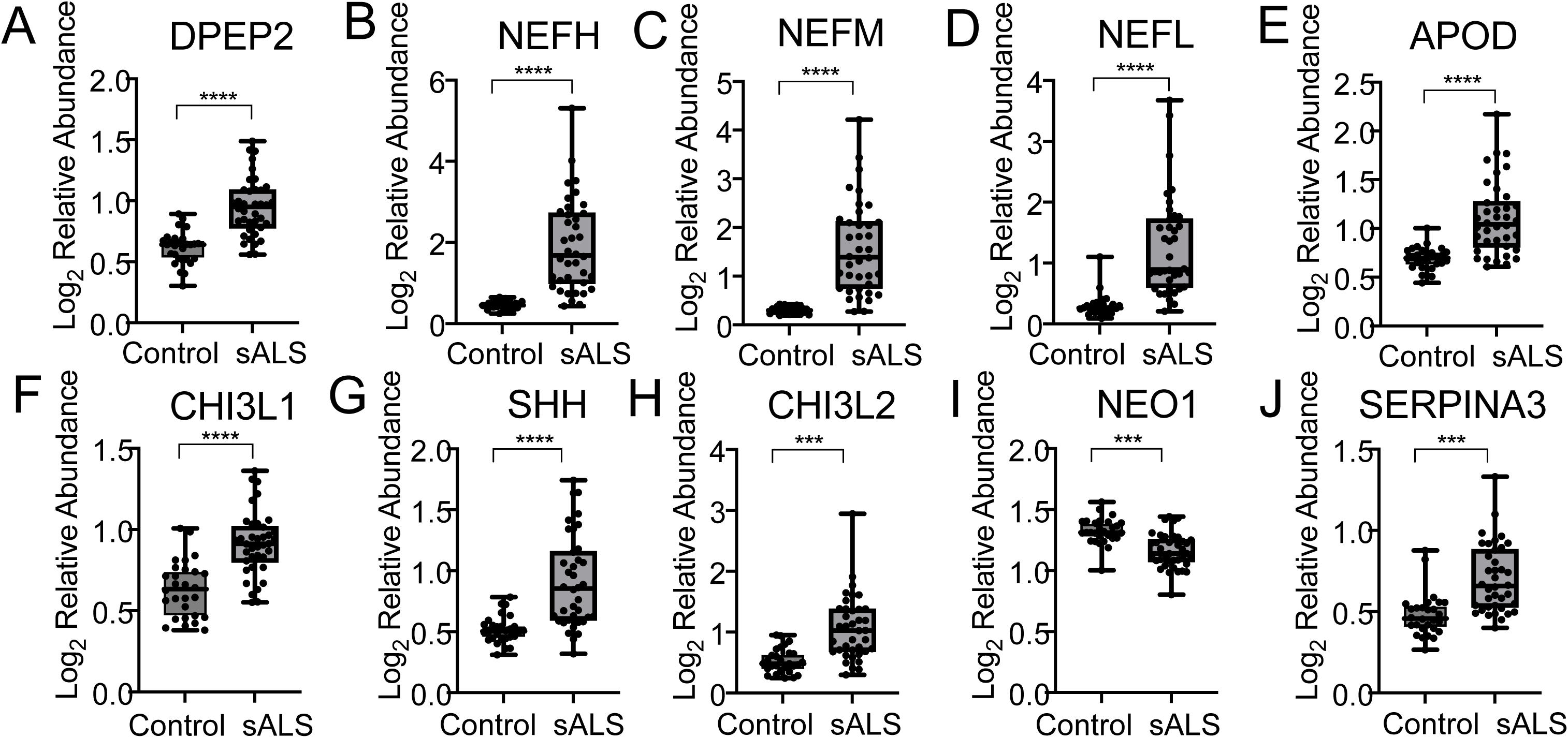
Top significant differentially expressed proteins in sALS CSF compared to control Box plots of the 10 most significant proteins showing log_2_ relative abundance in control compared to sALS CSF. Each dot represents relative protein abundance in a single sample and line inside each box represents the median. Whiskers reflect minimum and maximum values. Proteins are shown in order of highest to least significance and include, A) dipeptidase 2 (DPEP2), B) neurofilament heavy (NEFH), C) neurofilament medium (NEFM), D) neurofilament light (NEFL), E) apolipoprotein D (APOD), F) chitinase 3-like protein 1 (CHI3L1), G) sonic hedgehog (SHH), H) chitinase 3-like protein 2 (CHI3L2), I) neogenin 1 (NEO1), and J) alpha1-antichymotrypsin (SERPINA3). Benjamini-Hochberg corrected adjusted p-values denoted *, p < 0.05; **, p < 0.01; ***, p < 0.001, ****, p < 0.0001.

### Proteomic analysis of differentially expressed proteins in sALS plasma

We identified 1,118 proteins in plasma using the 35-plex TMTpro-MS approach (Figure 3 A) and included 646-762 proteins in each cohort after filtering for plasma samples that exhibited at least 50% quantified proteins (Supp. Figure 1 C-D). The filtering step excluded proteins only measured in healthy controls (n= 47) or sALS (n= 20) (Supp. Table 5). Plasma proteomic analysis included the same participants and dates of collection as analyzed for CSF as well as an additional three healthy controls and two sALS (HC: n= 31; sALS: n= 41). Six proteins were significantly increased and two decreased in sALS relative to healthy controls (Figure 3 B). In contrast to CSF, neurofilament proteins were not detected in plasma, consistent with other TMT-MS based studies ^29^. The most significantly increased DEPs include muscle-expressed proteins carbonic anhydrase 3 (CA3), myoglobin (MB), and creatine kinase, M-type (CKM), mediators of axon guidance, neuronal cell adhesion molecule (NRCAM), neural cell adhesion molecule-1 (NCAM1), and lipoprotein-associated phospholipase A2 (PLA2G7)^30^ which been shown to be active during inflammatory and oxidative stress conditions (Figure 3 C-F, H, J, Supp. Table 5). Contrarily, serpin family B member 6 (SERPINB6) and chondroadherin (CHAD) were decreased compared to control (Figure 3 G, I). Notably, NRCAM, CHAD, and PLA2G7 were also differentially expressed between control and sALS in CSF (Supp. Table 3) suggesting that plasma proteins altered in disease could reflect CNS dysregulation.

**Figure 3.**
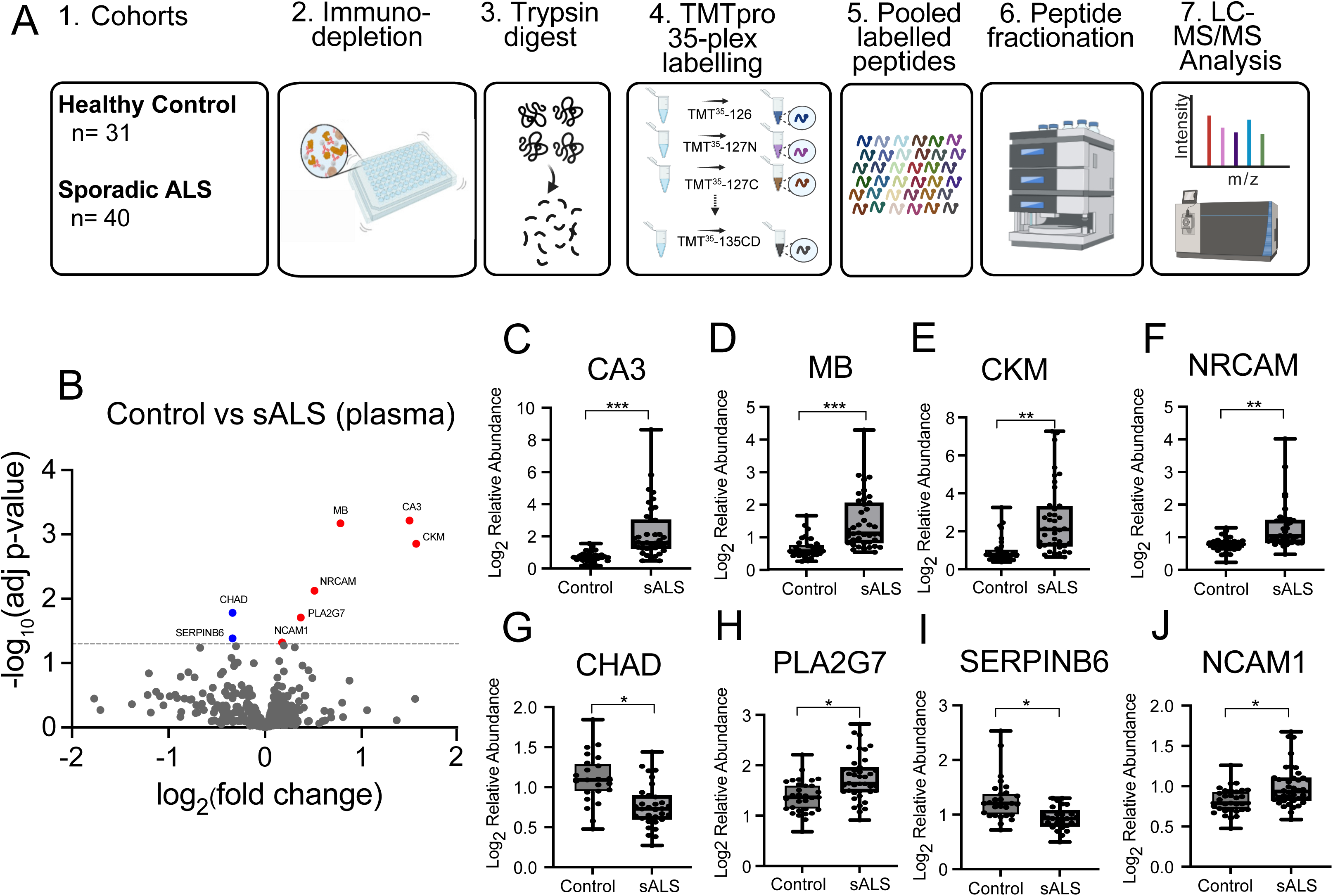
Overview of 35-plex TMTpro-MS methodology for plasma and differentially expressed proteins between control and sALS. Flow diagram of A) proteomic workflow used to determine proteomic differences in plasma from (1) control versus sporadic ALS (sALS). Plasma is (2) immune-depleted to remove abundant proteins, (3) enzymatically digested, (4) undergoes TMT-labelling, and (4) samples are pooled prior to (5) analysis by liquid chromatography tandem mass spectrometry (LC-MS/MS). Volcano plot showing differentially expressed plasma proteins between B) controls (n= 31) versus sALS (n= 40). Box plots of the 8 significant proteins showing log_2_ relative abundance in control compared to sALS plasma. Each dot represents relative protein abundance in a single sample and line inside each box represents the median. Whiskers reflect minimum and maximum values. Proteins are shown in order of highest to least significance and include, C) carbonic anhydrase 3 (CA3), D) myoglobin (MB), E) creatine kinase M-type (CKM), F) neuronal cell adhesion molecule (NRCAM), G) chondroadherin (CHAD), H) platelet-activating factor acetylhydrolase (PLA2G7), I) serpin B6 (SERPINB6), and J) neural cell adhesion molecule 1 (NCAM1). Benjamini-Hochberg corrected adjusted p-values denoted *, p < 0.05; **, p < 0.01; ***, p < 0.001.

### Comparison of 35plex TMTpro-MS to antibody-based proteomics

Advances in proteomic technologies have emerged to allow high-throughput, multi-plexed, and highly sensitive measures of protein in minute volumes of CSF or blood to facilitate biomarker discovery. However, cross-platform comparisons are needed to determine how they may agree with, complement, or differ from one another to aid appropriate selection.

We compared detection of significant DEPs by untargeted 35-plex TMTpro-MS and Olink Explore HT in CSF and plasma from healthy controls (TMTpro-MS: HC n= 28; Olink: HC n= 24) and sALS (TMTpro-MS: sALS n= 39; Olink: sALS n= 32). All samples analyzed by Olink HT were represented in the 35-plex TMTpro-MS cohorts. We measured 2,090 proteins in CSF using our unbiased 35plex TMTpro-MS workflow while Olink HT reports measures of 5,401 panel proteins, respectively, of which 1,060 proteins were detected by both methods (Supp. Table 8). Notably, each method measured distinct proteins not observed in the other with 1,030 proteins that were only detected by TMTpro-MS and 4,341 by Olink. We identified 40 significant DEPs in CSF using Olink HT (Figure 4 A-B) compared to 297 with TMTpro-MS. We identified 12 common hits including established biomarkers NEFL, CHI3L1, and CHIT1 as well as proteins with less defined links to disease, APOD, SERPINA3, IL1R2, CD48, PRSS2, CCL2, RBP4, FABP1, and REG1A (Supp. Table 9). However, TMTpro-MS identified 111 DEPs (e.g., NEFH, NEFM) not represented in the Olink protein library and an additional 174 DEPs (e.g., GPNMB, DPEP2, SHH) that were not significant based on adjusted p-value <0.05 thresholds in Olink HT analyses (Supp. Table 9). Alternatively, Olink identified 22 unique DEPs not represented by TMTpro-MS including IL-18, CCL3, CCL23, CCL15, as supported by gene ontology analyses suggesting enrichment of CSF chemokine/ cytokine pathways proteins using Olink HT (Supp. Table 7), as well as PRPH and HSPB6 which have previously been to be found to be elevated in ALS biofluids^12,31^ (Supp. Table 9).

**Figure 4.**
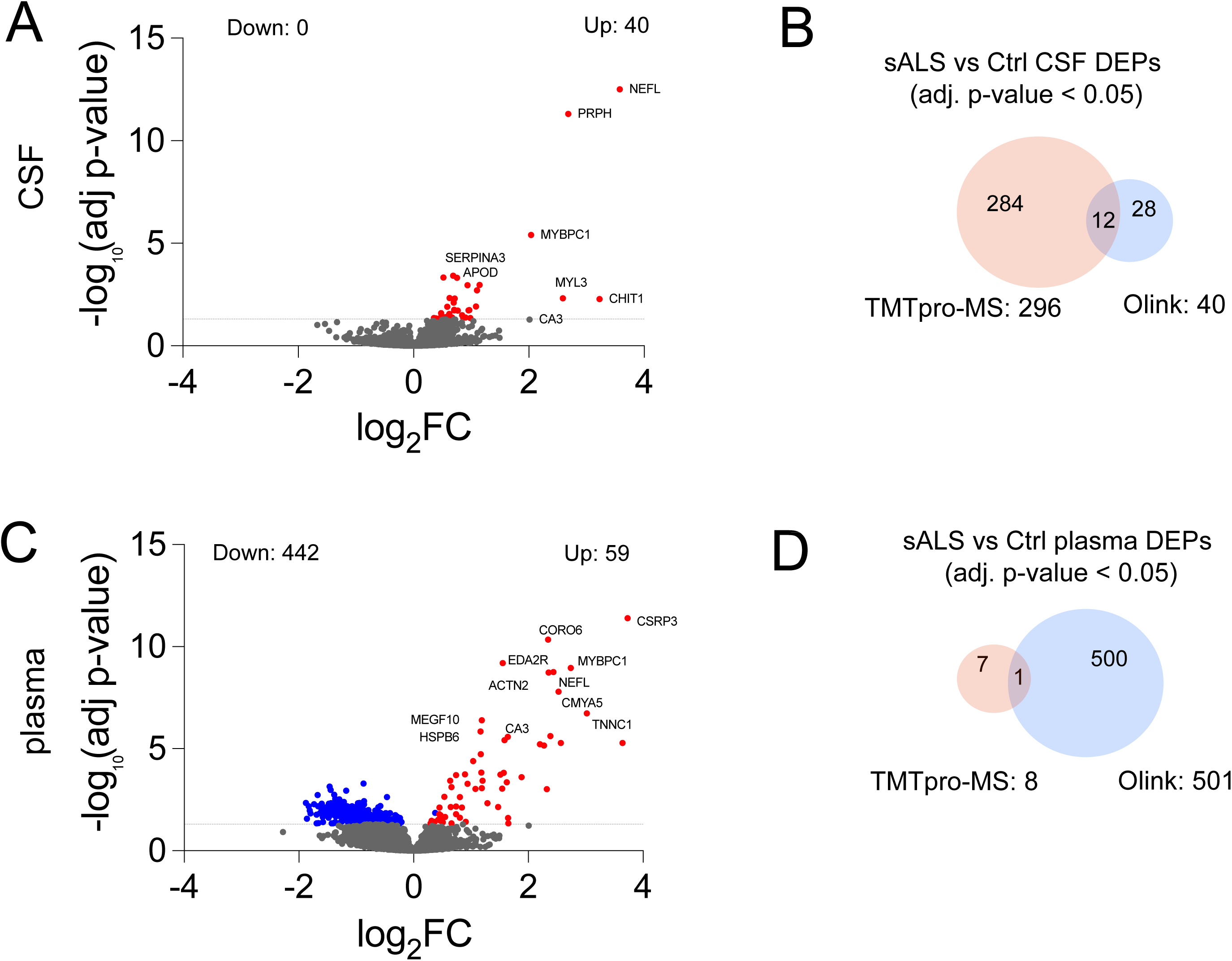
Differential expression analysis of ALS and control CSF and plasma using antibody-based proteomics and comparison to 35plex TMTpro-MS. Volcano plot showing A) differentially expressed CSF proteins between controls (n= 24) versus sALS (n= 32) using Olink HT. B) Venn diagram showing overlap of DEPs identified between healthy control and sALS in CSF by 35plex TMTpro-MS and Olink HT. Volcano plot showing C) differentially expressed plasma proteins between controls (n= 24) versus sALS (n= 32) using Olink HT. D) Venn diagram showing overlap of DEPs identified between healthy control and sALS in plasma by 35plex TMTpro-MS and Olink HT.

Plasma analysis (TMTpro-MS: HC n= 31, sALS n= 41; Olink: HC n= 24, sALS n= 32), detected 668 proteins with TMTpro-MS and measured 5,416 proteins using Olink platform with an overlap of 342 common proteins assayed (Supp. Table 10). We identified 326 proteins that were only detected by TMTpro-MS and not represented by the Olink HT platform. Differential expression analysis of control and sALS plasma by Olink HT uncovered 501 significant DEPs between HC and sALS in plasma (Figure 4 C). However, only one DEP was shared with 35-plex TMTpro-MS, carbonic anhydrase 3 (CA3) (Figure. 4 D, Supp. Table 11). The antibody-based proximity extension assay approach identified 474 unique DEPS not represented by TMTpro-MS protein library and an additional 26 DEPs that were not significant based on adjusted p-value <0.05 threshold in TMTpro-MS analysis. Conversely, TMTpro-MS identified two significant DEPs that were not represented in Olink panel, MB and CKM. We also identified five DEPs with unbiased mass spectrometry-based proteomics that were not significant using the Olink platform including NRCAM, CHAD, PLA2G7, SERPINB6, and NCAM1.

### Tissue enrichment of TMTpro-MS and Olink DEPs

In order to determine the tissue origin of differentially expressed CSF and plasma proteins determined by 35-plex TMTpro-MS and Olink, we performed tissue expression analysis of upregulated and downregulated CSF and plasma DEPs using gene ontology-based enrichment analysis. In CSF, upregulated DEPs from TMTpro-MS were predominantly enriched with immune/ blood-derived proteins (27.3%) followed by central nervous system (CNS) (16%), and muscle-derived proteins (15.3%) whereas Olink CSF DEPs showed equal enrichment of immune/ blood (30%) and muscle-derived proteins (30%) but lower proportion of CNS proteins (10%) (Supp Figure 2 A) suggesting that 35-plex TMTpro-MS is more sensitive for brain-derived proteins. Although no downregulated DEPs were detected by Olink, downregulated CSF DEPs identified by TMTpro-MS were likewise defined by CNS-derived proteins (46.6%) followed by muscle (9.5%) and immune/ blood-derived proteins (7.4%) (Supp Figure 2 B). Upregulated plasma DEPs from Olink were characterized by enrichment of muscle-derived proteins (54.2%) consistent with gene ontology analysis that suggest roles in myofibril assembly and function (Supp Figure 2 C, Supp Table 7). Olink downregulated DEPs were more equally represented by muscle (15.2%), immune/ blood (13.3%), and CNS-derived proteins (12.2%) (Supp Figure 2 D). These results highlight advantages of TMTpro-MS for detection of brain-derived proteins and alternatively, the superior sensitivity of Olink for muscle-derived proteins in CSF and plasma.

### Plasma proteome coverage of TMTpro-MS and Olink

In order to determine plasma proteome coverage of the two methods, we used estimated concentrations of a reference set of 4,889 secreted blood proteins derived from the Peptide Atlas, ww.peptideatlas.org) and Human Protein Atlas as described (HPA, www.proteinatlas.org) ^32,21^. UniProt IDs were used to match proteins between TMTpro-MS, Olink, and reference dataset. We were able to match 569 of 668 proteins detected in plasma by TMTpro-MS and 1525 of 5416 proteins detected in plasma by Olink, to estimated blood concentrations and plotted the distribution of estimated concentrations for all proteins detected by either method or detected solely by TMTpro-MS or Olink (Supp Figure 3 A-B). Of the 342 proteins that were detected in plasma by both platforms, we matched 330 with estimated concentrations. We found that TMTpro-MS demonstrated better coverage of moderate to high abundance proteins while Olink provided superior coverage of low abundance proteins as previously described ^21^. Our comparative analysis indicates that proteomic coverage provided by unbiased and antibody-based proteomics in ALS CSF and plasma are distinct and complementary.

### ELISA validation of 35plex TMTpro-MS hits

Although proteomic approaches have utility for unbiased biomarker discovery, orthogonal validation of candidate biomarkers is critical for confirmation and bridging these findings to a clinically accessible platform. We utilized commercially available enzyme-linked immunosorbent assays (ELISAs) to verify differentially expressed proteins identified by 35plex TMTpro-MS in CSF and plasma in an independent cohort of healthy control and ALS biofluids obtained from the Washington University ALS biorepository. We used ELISA to quantify CKM and CHAD, proteins found to be increased and decreased in ALS, respectively, based on plasma TMTpro-MS analyses, using plasma from healthy control (n= 10) and sALS (n= 20). Contrary to TMTpro-MS findings, the concentration of CHAD in plasma was no different between controls and sALS (Figure 5 A). However, CKM concentration was found to be increased in sALS plasma, providing secondary validation of CKM as a protein altered in disease (Figure 5 B). As SERPINA3 was identified as novel differentially increased protein in ALS CSF, we compared abundance of SERPINA3 in healthy control (n= 10) and sALS (n= 15) using ELISA and found that SERPINA3 was increased in ALS confirming findings from TMTpro-MS (Figure 5 C). As ELISAs are considerably more feasible to implement in lab-based and clinical setting, our ability to confirm TMTpro-MS plasma and CSF hits further aid to validate the platform and provide assessable approaches for further biomarker development.

**Figure 5.**
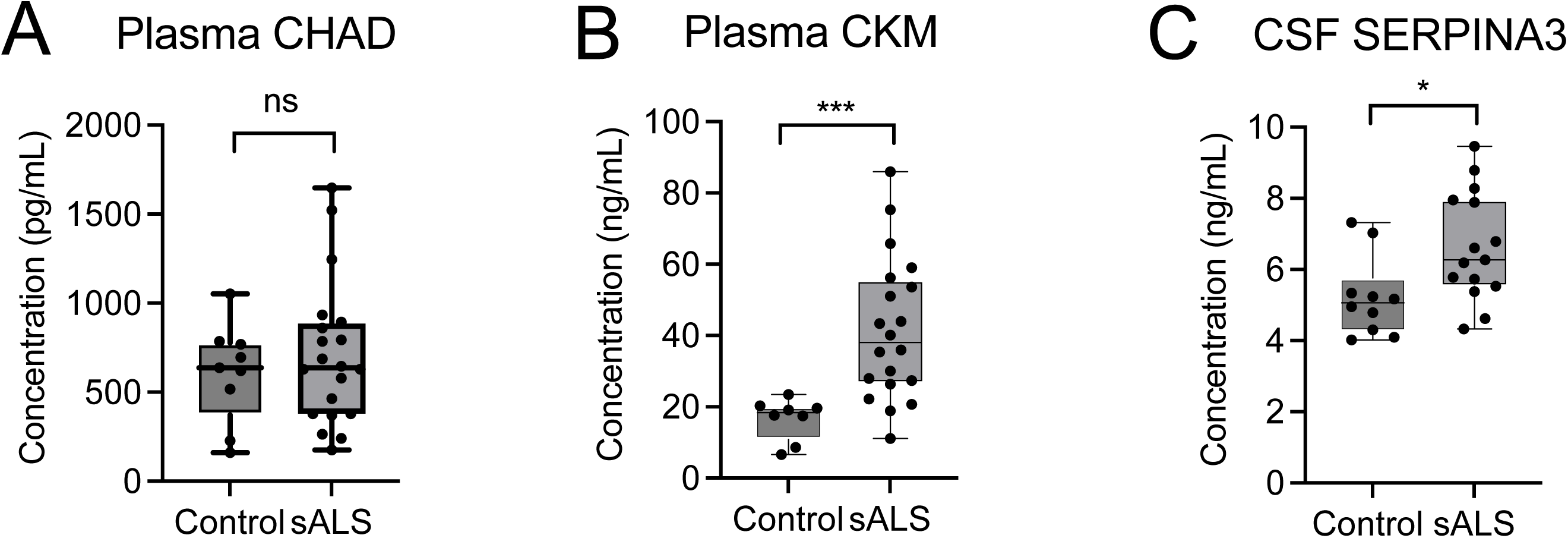
ELISA validation of 35plex TMTpro-MS hits ELISA was performed to provide orthogonal validation of putative plasma and CSF hits determined by 35plex TMTpro-MS using an independent biofluid cohort. A) Concentration of chondroadherin (CHAD) in healthy control and sALS plasma. B) Concentration of muscle creatine kinase (CKM) in healthy control and sALS plasma. C) Concentration of serpin family A group 3 (SERPINA3) in healthy control and sALS CSF. p-values denoted *, p < 0.05; ***, p < 0.001, ns- not significant.

## Conclusions

Reliable diagnostic, prognostic, and pharmacodynamic biomarkers are necessary to advance clinical care and therapeutic development for ALS, a rapidly progressive neurodegenerative condition that often evades diagnosis for nearly a year and for which disease-modifying therapies are lacking for a majority of patients. In this cross-sectional, cross-platform study, we performed proteomic profiling of CSF and plasma from sporadic ALS and controls and compared the performance of 35-plex TMTpro-MS to Olink proteomics.

Advances in tandem mass tag (TMT) multi-plexing approaches provide enhanced throughput and reduced variability amenable to analysis of large patient cohorts while allowing sensitive detection of proteoforms. Comparison of sALS and control CSF 35-plex TMT-MS proteomes revealed a number of previously established biomarkers including neurofilaments (NEFH, NEFM, NEFL), chitinases (CHI3L1, CHI3L2, CHIT1), and GPNMB among the most significant DEPs, indicating the robustness of the 35-plex TMTpro-based methodology. The most significant novel biomarker candidates included proteins involved in axon guidance and neuronal maintenance (i.e. SHH, NEO1), lipid metabolism (APOD), macrophage-mediated inflammation (DPEP2) ^33^, and serine protease inhibition (SERPINA3). Notably, APOD has been shown to be elevated in CSF from Alzheimer’s disease (AD) patients^34,35^ and SERPINA3 has been shown to be associated with amyloid plaques in postmortem AD brain^36^ indicating a neurodegenerative association for these proteins. Further studies are required to ascertain their role in disease and to validate them as potential biomarkers.

The accessibility and ease of collecting plasma is desirable for next-generation ALS clinical biomarkers to facilitate diagnosis, prognosis, and screening for clinical trial enrollment. However, whereas CSF provides a proxy of central nervous system alterations in disease, the plasma compartment more likely reflects changes in the periphery. Plasma TMTpro-MS analysis of healthy control and sALS revealed six increased DEPs, CA3, MB, CKM, NRCAM, PLA2G7, and NCAM1, and two decreased DEPs, CHAD and SERPINB6. CA3, MB, and CKM are muscle -enriched proteins reported to be increased in ALS plasma^12^. Notably, neurofilaments were not detected by 35-plex TMTpro-MS similar to other reports^29^, likely due to relative complexity of plasma compared to CSF. NRCAM, NCAM1, SERPINB6 and CHAD have not previously been identified as differentially expressed in ALS plasma. Notably, NRCAM, SERPINB6, and CHAD were also found to be significant differentially expressed proteins in CSF. NCAM1 is a neuronally-expressed cell adhesion molecule involved in axon guidance that has been detected in surviving motor neurons of SOD1 G93A mice but has not been established in human disease ^37^. PLA2G7 is a lipoprotein-associated phospholipase active during inflammatory and oxidative stress conditions and has been identified as a risk factor for dementia ^30,38^ and found to be increased in ALS plasma ^12,39^ but remains understudied. Downregulated plasma protein CHAD encodes chondroadherin which regulates bone and cartilage integrity ^40^ while SERPINB6 is a serine protease inhibitor found to associate with neurofilament aggregates in ALS motor neurons ^41^. These findings highlight the potential for this approach to identify candidate plasma biomarkers implicated in multiple disease pathways.

Mass spectrometry facilitates unbiased biomarker discovery but require orthogonal validation using independent platforms and cohorts to determine biological relevance and absolute quantitation of putative biomarkers. We used commercial immunoassays and a validation plasma cohort to validate two plasma proteins identified by 35plex TMTpro-MS as decreased (CHAD) and increased (CKM) in plasma and determined that although CHAD levels were no different between sALS and control, CKM was confirmed to be elevated. Similarly, we confirmed in a separate CSF cohort that SERPINA3 was increased in the CSF of ALS patients by ELISA. As mass spectrometry requires specialized expertise and high entry costs, establishing alternative assays for putative biomarker hits is important to improve accessibility and lower barriers to clinical translation.

Beyond mass spectrometry, numerous improvements have been made in proteomic approaches to screen for candidate disease biomarkers. Multiple technologies allow high-throughput, multi-plexed, and highly sensitive measures of protein in minute volumes of CSF or blood to facilitate clinical workflows for biomarker discovery. These modalities rely on unique proprietary detection strategies involving DNA aptamers (i.e., SomaScan), antibody recognition with nucleic acid amplification (i.e., NULISA), or proximity extension assays (i.e., Olink) ^42^. We performed parallel proteomic analyses using TMTpro-MS and Olink HT on matching CSF and plasma from sALS and controls and compared the performance of the two methodologies. In CSF, TMTpro-MS showed superior sensitivity over Olink, identifying 297 DEPs compared to 40 DEPs by Olink HT and 20 DEPs with SomaScan 7K in a prior study ^39^. There were 12 DEPSs showing significance in both platforms, including known ALS biomarkers, NEFL, CHIT1, and CHI3L1. Notably, TMTpro-MS detected the full spectrum of neurofilaments in CSF including NEFM, and NEFH as well as neuropentraxin-2 (NPTX2), a secreted neuronal protein whose expression is regulated by TDP-43 binding and linked to shorter survival in ALS^43^. PRPH, a subunit complexed with neurofilaments in peripheral nerve was only detected by Olink, indicative of its lower abundance defined by established intermediate filament stoichiometries ^44^ while NEFM and NEFH were not incorporated in the Olink panel and therefore not detected. In plasma, Olink showed superior sensitivity detecting 501 DEPs compared to only 8 by TMTpro-MS. The only DEP shared between the two platforms in plasma was CA3. Plasma is a complex matrix and TMTpro-MS analysis has traditionally been limited by ratio compression due to co-elution of low and high abundance peptides resulting in signal suppression of the former. The plasma profile detected by Olink was largely driven by skeletal muscle-enriched proteins (e.g., MYBPC1, TNNC1, CSRP3), suggesting a systemic proteomic footprint defined by muscle-derived proteins and consistent with prior reports employing aptamer and antibody-based proteomic technologies^12,39^.

Comparison of 35-plex TMTpro-MS and Olink revealed how the methods complement one another in ALS biomarker development. Olink has superior detection of low abundance proteins and better discrimination of DEPs in plasma, while 35-plex TMTpro-MS showed superior sensitivity in CSF for identifying potential ALS biomarkers. Mass spectrometry affords intrinsic peptide and protein-level specificity, higher sensitivity in CSF, and potential to identify post-translationally modified and alternatively spliced proteoforms including cryptic peptide species that derive from TDP-43 loss-of-function^18,45^. These strengths are beneficial for examining disease pathomechanisms in light of aberrant oxidative/ proteostatic stress and predominant TDP-43 pathology in ALS and identifying pathway-specific disease biomarkers. Alternatively, the superior sensitivity and high-throughput capacity of antibody-based technologies for plasma proteins suggest technologies such as Olink may be more amenable to broad screening of disease cohorts. Our initial unbiased cross-sectional multi-omic analysis of biofluids from the Target ALS Global Natural History Study serves as a foundation for biomarker and therapeutic discovery in ALS and open resource for the broader research community. These studies provide the basis of a standardized methodology for future analyses of longitudinal biofluids from internationally diverse ALS cohorts.

## Data Availability

All data produced in the present study are available upon reasonable request to the authors

https://dataengine.targetals.org/collections

## Acknowledgements

The authors are grateful to the participants of the TALS GNHS as well as their families who contributed specimens. This study would not have been possible without their support and desire to advance ALS research. We acknowledge the support of Target ALS for funding this work. HZ is a Wallenberg Scholar and a Distinguished Professor at the Swedish Research Council supported by grants from the Swedish Research Council (#2023-00356, #2022-01018 and #2019-02397), the European Union’s Horizon Europe research and innovation programme under grant agreement No 101053962, Swedish State Support for Clinical Research (#ALFGBG-71320), the Alzheimer Drug Discovery Foundation (ADDF), USA (#201809-2016862), the AD Strategic Fund and the Alzheimer’s Association (#ADSF-21-831376-C, #ADSF-21-831381-C, #ADSF-21-831377-C, and #ADSF-24-1284328-C), the European Partnership on Metrology, co-financed from the European Union’s Horizon Europe Research and Innovation Programme and by the Participating States (NEuroBioStand, #22HLT07), the Bluefield Project, Cure Alzheimer’s Fund, the Olav Thon Foundation, the Erling-Persson Family Foundation, Familjen Rönströms Stiftelse, Familjen Beiglers Stiftelse, Stiftelsen för Gamla Tjänarinnor, Hjärnfonden, Sweden (#FO2022-0270), the European Union’s Horizon 2020 research and innovation programme under the Marie Skłodowska-Curie grant agreement No 860197 (MIRIADE), the European Union Joint Programme – Neurodegenerative Disease Research (JPND2021-00694), the National Institute for Health and Care Research University College London Hospitals Biomedical Research Centre, the UK Dementia Research Institute at UCL (UKDRI-1003), and an anonymous donor. RB was supported by grants from the Target ALS Foundation (CF-2026-PMT-S2 and CF-2026-BFC-S2), NIH/NINDS grant OT2 NS136939-03, and Department of Defense grants (AL2402-49, AL2201-64, AL2201-03, and AL2000-61). CVL was supported by the Target ALS Foundation and NIH/NINDS grant R01 NS138499

## Author contributions

LD and DW were involved in study design, data collection, and data curation. DY was involved in analysis of clinical data, processed proteomic data and wrote the manuscript. JG and HZ were involved in study design, data analysis, technical oversight, and intellectual guidance. SW conducted 35-plex TMT-MSpro proteomics studies. LG performed immunoassay studies. HT, KN, GG, BH, LN, WD, CH, JL, and SN were involved in data collection. BO, TMM, JR, MBH, NS, SL, NS, BH were involved in data collection and clinical site supervision. MR was involved in study conception. AE was involved in study conception and study design. RB was involved in study conception, study design, technical guidance, and intellectual guidance. CVL was involved in study design, data analysis, intellectual guidance, and wrote the manuscript. All authors reviewed the manuscript.

## Materials and Correspondence

## Supplementary Figures

Supplementary Figure 1. CSF and plasma protein detection by 35plex TMTpro-MS

For CSF, a total of 2,875 proteins across 14 TMT sets were detected (A). After quality control to include CSF samples with at least 50% of proteins having quantification, 2,120 and 2,163 proteins among our sALS and healthy control cohorts, respectively were included for comparative analysis (B). For plasma, a total of 1,118 proteins were detected (C). Removel of proteins absent in at least 50% of cases yielded 688 and 715 plasma proteins that were compared between sALS and control, respectively.

Supplementary Figure 2. Tissue enrichment of CSF and plasma differentially expressed proteins (DEPS) in sALS.

DEPs were assigned to tissue-associated functional categories based on Gene Ontology Biological Process (GO BP) annotations and displayed as platform-specific proportions (100% stacked bars; 35-plex TMTpro-MS and Olink HT), with total DEP counts indicated above each bar. Upregulated and downregulated DEPs in CSF are shown in (A) and (B), and upregulated and downregulated DEPs in plasma in (C) and (D), respectively. The Olink HT platform predominantly captured muscle/atrophy-associated proteins across both biofluids, while TMTpro-MS captured a substantial proportion of CNS-associated proteins in CSF (46.6% of downregulated and 16.0% of upregulated CSF DEPs).

Supplementary Figure 3. Plasma proteome coverage of 35plex TMTpro-MS versus Olink HT

Kernel density estimate (KDE) plots matching estimated plasma protein concentrations ^32^ to A) all plasma proteins detected in TMTpro-MS and Olink and B) plasma proteins detected by only one method. Differences in protein concentration between platforms were tested using a two-sided Wilcoxon rank-sum test.

## Supplementary Tables

Supp Table 1. Target ALS CSF cohort.

ALSFRS-R- amyotrophic lateral sclerosis functional rating score-revised, STD- standard deviation

Supp Table 2. Target ALS plasma cohort.

ALSFRS-R- amyotrophic lateral sclerosis functional rating score-revised, STD- standard deviation, N/A- not available

Supp Table 3. CSF Proteomic Comparisons for 35-plex TMT-MS and Olink (full list).

HC- healthy control, sALS- sporadic amyotrophic lateral sclerosis, STD- standard deviation, CV-coefficient of variation, adj FDR- adjusted false discovery rate, log2FC- log2 fold-change

Supp Table 4. Plasma Proteomic Comparisons for 35-plex TMT-MS and Olink (full list).

HC- healthy control, sALS- sporadic amyotrophic lateral sclerosis, STD- standard deviation, CV-coefficient of variation, adj FDR- adjusted false discovery rate, log2FC- log2 fold-change

Supp Table 5. List of proteins detected in TMTpro-MS, list of proteins detected in only control or sALS.

TMTpro-MS- 35-plex isobutyl proline tandem mass tag mass spectrometry, HC- healthy control, sALS- sporadic amyotrophic lateral sclerosis CSF- cerebrospinal fluid

Supp Table 6. CSF and plasma DEPs identified by 35plex TMTpro-MS analyses

DEPs- differentially expressed proteins, TMTpro-MS- 35-plex isobutyl proline tandem mass tag mass spectrometry, HC- healthy control, sALS- sporadic amyotrophic lateral sclerosis CSF-cerebrospinal fluid, adj FDR- adjusted false discovery rate, log2FC- log2 fold-change

Supp Table 7. Gene ontology analysis of CSF and plasma DEPs identified by 35plex TMT-MS analysis.

DEPs- differentially expressed proteins, CSF- cerebrospinal fluid, HC- healthy control, sALS-sporadic amyotrophic lateral sclerosis

Supp Table 8. Common and unique CSF proteins detected by 35plex TMTpro-MS and Olink HT. CSF- cerebrospinal fluid, HC- healthy control, sALS- sporadic amyotrophic lateral sclerosis, TMTpro-MS- 35-plex isobutyl proline tandem mass tag mass spectrometry

Supp Table 9. Common and unique CSF DEPs detected by 35plex TMTpro-MS and Olink HT.

DEPs- differentially expressed proteins, CSF- cerebrospinal fluid, HC- healthy control, sALS-sporadic amyotrophic lateral sclerosis, TMTpro-MS- 35-plex isobutyl proline tandem mass tag mass spectrometry

Supp Table 10. Common and unique plasma proteins detected by 35plex TMTpro-MS and Olink HT

HC- healthy control, sALS- sporadic amyotrophic lateral sclerosis, TMTpro-MS- 35-plex isobutyl proline tandem mass tag mass spectrometry

Supp Table 11. Common and unique plasma DEPs detected by 35plex TMTpro-MS and Olink HT

DEPs- differentially expressed proteins, HC- healthy control, sALS- sporadic amyotrophic lateral sclerosis, TMTpro-MS- 35-plex isobutyl proline tandem mass tag mass spectrometry

Supp Table 12. WU CSF validation cohort

CSF- cerebrospinal fluid, STD- standard deviation

Supp Table 13. WU plasma validation cohort STD- standard deviation

