## Supplemental Figures 1-3 for "The Target ALS Global Natural History Study: Cross-platform proteomics to accelerate biofluid biomarker and drug target discovery in amyotrophic lateral sclerosis"

A

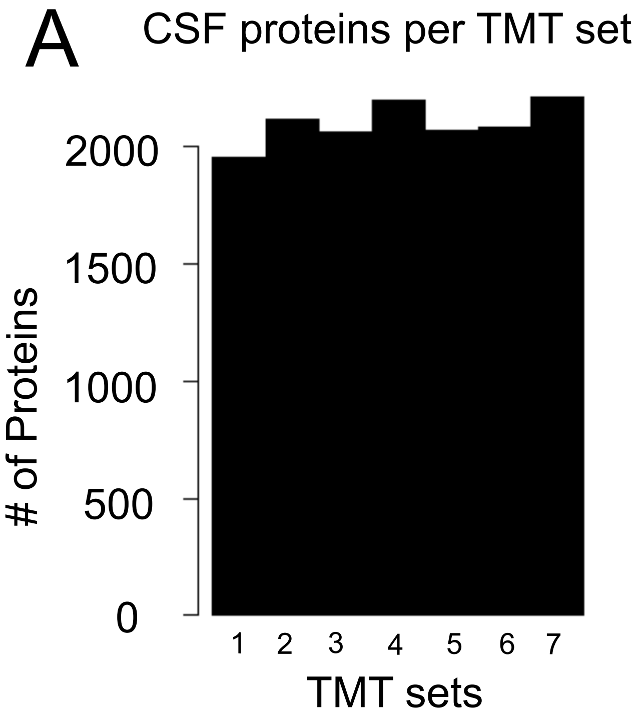


B

| | CSFprotein total | CSFprotein included | CSFprotein excluded |
|---|---|---|---|
| HC | 2875 | 2163 | 712 |
| sALS | 2875 | 2120 | 755 |

C

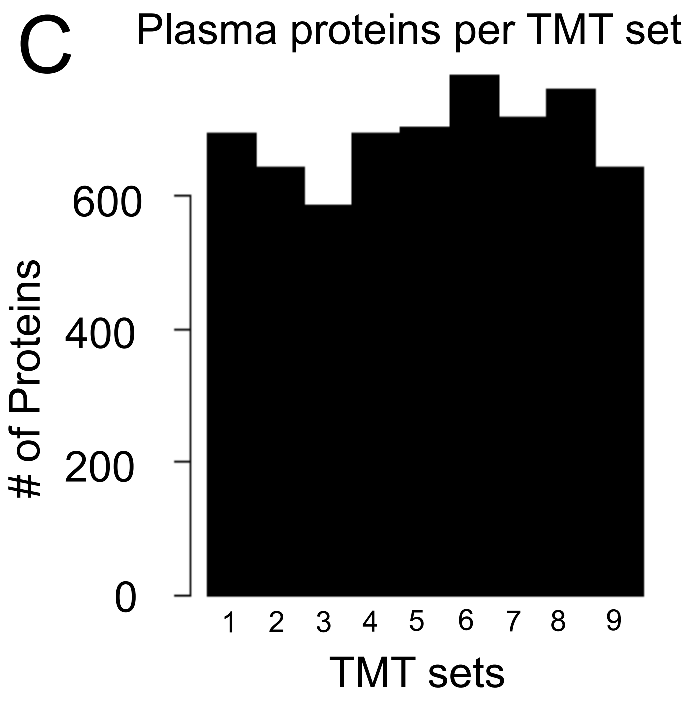


D

| | Plasma protein total | Plasma protein included | Plasma protein excluded |
|---|---|---|---|
| HC | 1118 | 715 | 403 |
| sALS | 1118 | 688 | 430 |

Supp Figure 1

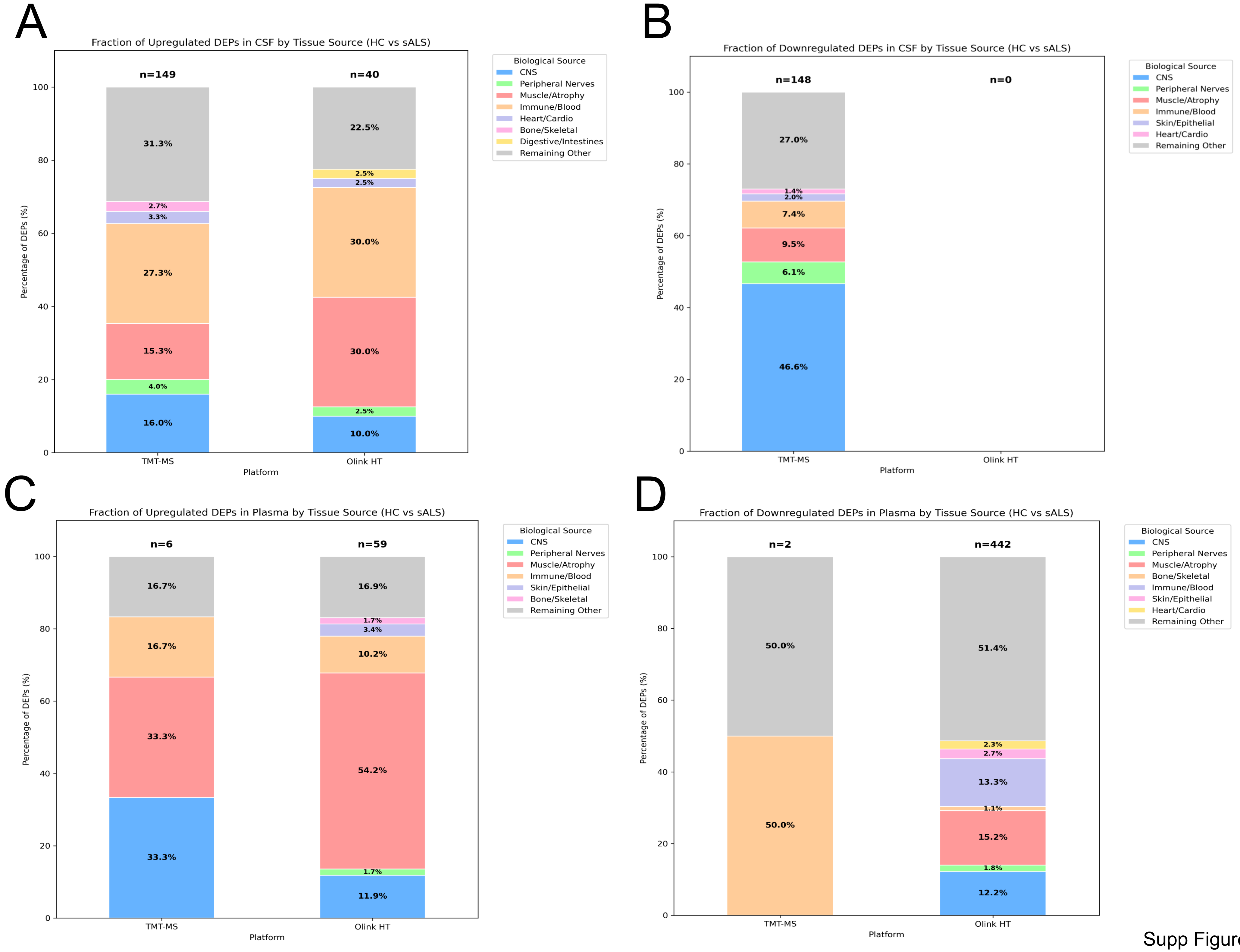


Supp Figure 2

A

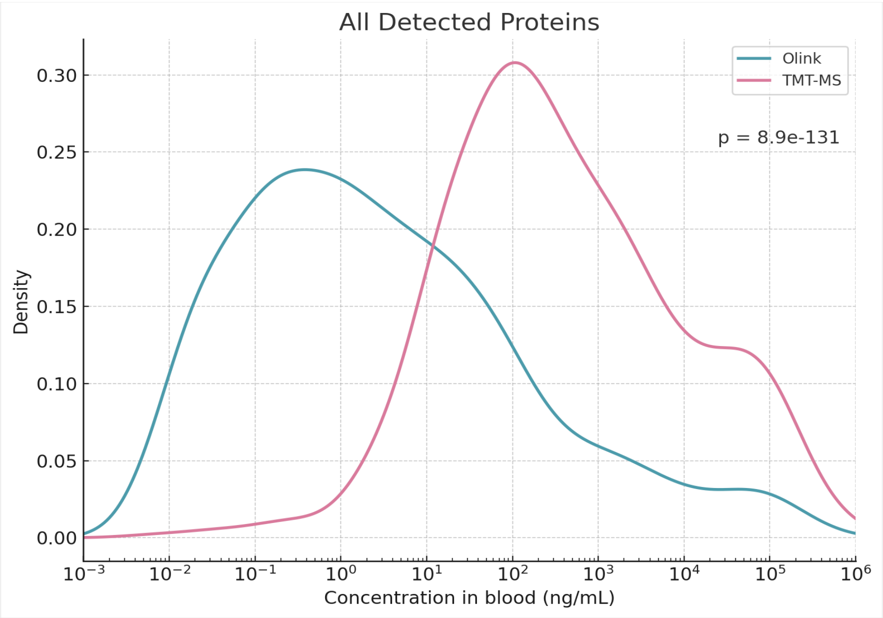


B

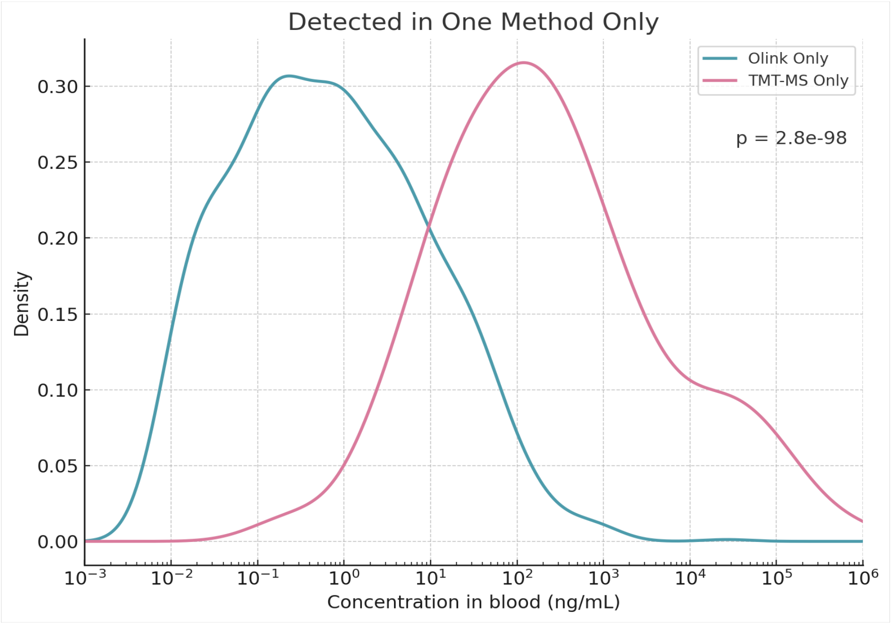


Supp Figure 3
